# Social, Physical Environmental and Organizational Factors related to Recreational Activity of Residents with Dementia in Long-Term Care Homes: A Scoping Review Protocol

**DOI:** 10.1101/2024.09.10.24312854

**Authors:** Ziying Zhang, Habib Chaudhury

## Abstract

**Introduction:** Recreational activity is a rising topic in long-term care settings as it contributes to residents’ physical and emotional wellbeing. As residents’ abilities of sustaining and initiating activities decline, the care environment becomes vital in supporting residents maintain meaningful engagement in activities and life. Understanding how various aspects of the care environment influence residents’ opportunities and quality of recreational activity engagement is a timely and relevant topic in the context of improving quality of life for residents with dementia.

The research questions guiding this scoping review are: (1) How do staff characteristics and organisational policies influence residents’ levels of participation in planned and self-directed activities? (2) What is the role of the physical environmental features in common spaces of the care setting in supporting residents’ activity participation?

**Methods and analysis:** This review will follow the Arksey and O’Malley scoping review methodology. The search strategy will be applied to five bibliographic and citation databases. Study selection will occur in two steps: first, two reviewers will screen the titles and abstracts of all search results, and second, the first author will independently conduct a full-text review. Data will be extracted from the included studies and analyzed using Braun and Clarke’s thematic analysis. The extracted data will be presented in a narrative format, accompanied by tables that reflect the review’s objectives.

**Ethics and dissemination:** Since the methodology of the study involves collecting data from publicly available publications, it does not require ethics approval. The findings will offer valuable insights to inform the design, practice and research of long-term care and recreational activities. The scoping review results will be disseminated through an open-access publication in a peer-reviewed journal.

**Strengths and Limitations:**

- This review offers a targeted view of environmental aspects related to the recreational activity of residents with dementia in long-term care settings. Evidence of both physical and social dimensions of the care environment related to recreational activities will be extracted and summarized.
- The review takes an inclusive approach to defining activities. Therefore, the findings will synthesize not only environmental factors that impact programmed recreational activities, but also those that encourage or limit opportunities for various kinds of meaningful spontaneous engagement.
- Being a scoping rather than a systematic review, the quality of the evidence will not be evaluated. The results and recommendations of scoping reviews cannot be graded since methodological appraisal of the quality of included studies will not be conducted.
- The review will be limited to English language studies.
- Review will only include studies based in long-term care homes, studies evaluated the recreational activities in adult day care centres, hospitals or other healthcare settings will not be captured.

## Introduction

With the growing aging population, a large and growing number of people with dementia are being cared for in long-term care (LTC) homes, which provide 24 hours, 7 days a week supervised care, including professional health services, personal care, and other services like meals, laundry, and housekeeping (British Columbia Ministry of Health, 2024). Enhancing the quality of life for people living with dementia has become a focal point and is acknowledged in the Canada’s first national strategy on dementia (Public Health Agency of Canada, 2019). Quality of life in dementia is a multidimensional concept, which involves physical and psychological wellbeing, social interaction, and positive/negative affect (Klapwijk et al., 2016).

Recreational activities, defined as discretionary pursuits with the purposes of entertainment, exercise, cognitive stimulation, creative expression, and socialization (Leitner & Leitner, 2012), are recognized as offering more opportunities for LTC residents to thrive than other types of activities, such as personal care (Fortune & Dupuis, 2018; Kelly, 2010; Phinney & Moody, 2011). Activity participation can improve the quality of life and well-being for residents with dementia (Allen, 2014; Marshall & Hutchinson, 2001; Tierney & Beattie, 2020), in terms of utilization of remaining cognitive and physical abilities (Chung, 2004; Edvardsson et al., 2014; Holthe et al., 2007; Palacios-Ceña et al., 2016), staying engaged in daily life and social interactions (Roland & Chappell, 2015), experiencing positive emotions (Beerens et al., 2016; Holthe et al., 2007; Kolanowski et al., 2001; Schreiner et al., 2004; Smit et al., 2016), maintaining autonomy and supporting personhood (Phinney et al., 2007), as well as mitigating the adverse effects of institutional living (Buettner & Fitzsimmons, 2002; Lopez & Dupuis, 2014; Tierney et al., 2022).

Despite these benefits, research revealed that residents’ needs for leisure are unfulfilled, spending most of their time unengaged or engaged in activities that do not meet their interests (Palacios-Ceña et al., 2016; Smit et al., 2014; Wood et al., 2005). The residents’ inactivity could magnify neuropsychiatric and behavioural symptoms, such as agitation, aggression, and apathy (Buettner & Fitzsimmons, 2003; MacDonald, 2006).

The challenges related to activity engagement are primarily attributed to three key factors: residents’ personal characteristics, the content of activities, and the institutional environment (Buettner & Fitzsimmons, 2003; Harmer & Orrell, 2008; Holthe et al., 2007; Smit et al., 2016; Tak et al., 2015). Although a fair amount of research has been conducted addressing the first two factors, research on environmental influences is still in a very early stage, especially those specifically address rather than general wellbeing and activities of daily lives.

Environmental and systemic barriers in the LTC homes hinder meaningful engagement. Fortune and Dupuis (2018) indicate that policy and practice in the LTC system has not yet shrugged off the long-standing dominance of biomedical model and implement a biopsychosocial approach entirely. Staff put focus on residents’ physical deficits and presentation of dementia, rather than their less overt psychosocial needs (Barbosa et al., 2014). Activities are often situated at the lower end of the power hierarchy in care homes, with little priority in activity programming, scarce personnel and limited financial resources (Ducak et al., 2016; Fortune & Dupuis, 2018). According to staff, policies and care plans do not fully appreciate the value of activity and consider providing activities as merely meeting regulatory requirements (Holthe, 2007). Additionally, a preference on quantitative results in the reporting system further prohibits staff from providing activities that people desire and/or have meaningful engagement (Misiorski & Kahn, 2006). Beyond the systemic barriers that staff may face, their skills of providing activities and knowledge about dementia also influence the provision of person-centered activities (Harmer & Orrell, 2008).

In addition to the social factors, the physical environment of care settings has been identified in several studies to relate to the residents’ engagement in recreational activities (Cohen-Mansfield et al., 2010; Holthe et al., 2007; MacDonald, 2006). Research indicates that the layout of the care unit (i.e., accessibility and variety in activity spaces), interior design can significantly impact residents’ opportunities and quality of activity engagement (Holthe et al., 2007; MacDonald, 2006; Smit et al., 2014; Voelkl et al., 2003). Besides, meaningful engagement in recreational activities further necessitates an enriched care environment that offers positive, high-quality environmental stimuli (Calkins, 2009; Cohen-Mansfield & Werner, 1999). However, overall, this body of knowledge, specifically addressing the impact of the physical environment on activities, remains underdeveloped, highlighting the need for a more in-depth and nuanced evidence base to guide architectural design and activity programming.

A growing number of studies have highlighted that care environment should be conceptualized as multiple interrelated components, which include physical environment (physical infrastructure, spatial layout, interior design, furniture, etc.), social environment (staffing model, staff skills etc.) and organizational aspects (philosophy of care, training and evaluation, etc.) (Seetharaman et al., 2022; Fleming & Purandare, 2010; Narsakka et al., 2022). Also, it has been noted that the effectiveness of physical environment is influenced by the staff/volunteer engagement and organisational policies/practices (Narsakka et al., 2022; Wood et al., 2005). For example, restricted access to a unit-based kitchen limits self-initiated activities (Saperstein et al., 2004). Therefore, examination of physical and social environmental and organizational aspects and their interrelationship would help gain a comprehensive understanding of the recreational activity experiences of residents with dementia in long-term care (Narsakka et al., 2021).

There has not been a review examining the enablers and barriers in the long-term care setting that affect participation in and experiences of recreational activities for residents with dementia. Two review studies have been conducted to understand the care environmental aspects related to the physical activity of residents with dementia in long-term care settings (Anderiesen et al., 2014; Narsakka et al., 2022). Findings from those studies highlighted various environmental features in the physical (safety, accessibility, home-likeness, etc.), social (supportive professionals, role of families, etc.), and organizational (policies, values, etc.) domains, as well as the interrelatedness of these features across dimensions. Given that the scope and nature of physical activities in these studies differ from those of recreational activities, the results from the two reviews are not transferable and cannot directly inform the design of the care environment or care practices.

As there is a growing consensus on the significance of recreational activity to residents’ quality of life, a comprehensive understanding of the influence of care setting on residents’ recreational activities participation and experience is important. This scoping review aims to: 1) review the existing literature examining the environmental characteristics and influence of long-term care homes on residents’ recreational activities; 2) understand the interrelationship of these factors, and 3) identify gaps for future research. Since the evidence in the area are relatively dispersed, a scoping review that systematically maps of the environmental aspects related to the recreational activities of residents is well-needed. Furthermore, findings from the review can provide evidence-based guidance in this expanding area of long-term care service provision.

## Methods

The scoping review method is adopted in order to most broadly capture the current discourse on this topic and point out gaps in the existing literature. To assist in the development of the protocol for the scoping review, Arksey and O’Malley’s (2005) framework and have incorporated the improvements put forth by Levac and colleagues (2010) to improve the rigour and consistency of the process. This framework proposes five mandatory stages which are outlined below. An optional sixth stage (consultation with stakeholders) is proposed, but our current study may not examine this due to the nascent stage of the project. Detailed process of each stage will be discussed below.

### Stage 1: Identifying the Research Questions

In order to guide the scoping study, broad research questions were developed, and a clearly articulated scope of inquiry was outlined.

How do the social and physical environment of the long-term care home facilitate or limit residents’ activities?

- How do activity programming, staff characteristics and organisational policies influence residents’ levels of participation in planned and self-directed activities?
- What is the role of the physical environmental features of the care setting in supporting residents’ activity participation?

### Stage 2: Identifying Relevant Studies

To ensure a comprehensive search for primary studies and other relevant literature, the team adopts an iterative approach to progressively refine the eligibility criteria, select databases, and determine the key search terms to use.

#### Eligibility criteria

The following inclusion and exclusion criteria will guide the development of the search strategy and be used to screen studies for inclusion in the review:

Inclusion Criteria:

1. Focused on residents with dementia in long-term care settings providing full-time care.
2. Took place in either the special care unit or regular care unit in the long-term care homes provided that residents with dementia or their families or staff were identified as primary participants of the research.
3. Investigated environmental aspects (physical or social) in relation to the recreational activity (programmed group recreational activities, one-one activities, household activities, and spontaneous recreational activities etc.) of residents with dementia in long-term care settings.
4. Were empirical studies.
5. Were published as research articles in scientific peer-reviewed journals in the English language.

Exclusion Criteria:

1. Involved community-dwelling older individuals, hospital patients, or residents living in facilities where full-time care was not provided.
2. Focused solely on individual factors or the contents of recreational activities.
3. Focused on physical activity or activities of daily living, self-care, or functional ability.
4. Were presentation, reviews or PhD dissertations.

#### Databases

The following five databases have been identified for searching for published studies: MEDLINE (Ovid), the Cumulative Index to Nursing and Allied Health Literature (CINAHL) (EBSCO), PsycINFO (ProQuest), AgeLine (EBSCO), Web of Science (Thompson Reuters). A limited Google Scholar search will be done to identify studies not retrieved by the databases. Hand searching of reference lists and forward citations from included studies will also be performed.

#### Search strategies

A pilot search will be conducted of the MEDLINE and CINAHL databases to identify additional relevant keywords and subject headings. The search is limited to English language papers and publications dates of 2000 to date of the search. Keyword search strings are outlined below.

### Long-term care

#### Keyword String

(“long-term care” OR “nursing home” OR “residential care” OR “assisted living” OR “special care unit”)

### Dementia

#### Keyword String

(“dementia” OR “Alzheimer’s” OR “cognitive impairment” OR “memory loss”)

### Activities

#### Keyword String

(“leisure” OR “recreational activities” OR “occupation” OR “engagement”)

### Environment

#### Keyword String

(“environment” OR “staff” OR “policy” OR “design” OR “culture” OR “practice” OR “practitioner”)

### Stage 3: Study Selection

Covidence literature review management software (Veritas Health Innovation Ltd, Melbourne, Australia) will be used to facilitate the screening process. After retrieved references are uploaded into Covidence, duplicate items will be removed by the software, with the authors manually checking for missed duplicates. Study selection will occur in a two-step process. First, the titles and abstracts of all papers retrieved by the searches will be screened by individual reviewers on the team, to determine eligibility based on the inclusion and exclusion criteria. In order to ensure the screening criteria are being equally applied, the two reviewers (ZZ and WW^1^) will independently assess the first 50 articles and then meet to discuss their conclusions. The second stage of the process will involve a full-text examination of sources for eligibility by the leading author (ZZ). An adapted PRISMA 2020 flow diagram will be used to graphically depict the article selection process and enhance the transparency about the searching and screening process. Discrepancies at each screening stage will be discussed in the three-author (ZZ, HC, and WW) meeting until consensus was reached.

### Stage 4: Charting the Data

During this stage, data will be extracted from the included studies, using a data extraction form jointly developed by the two authors (ZZ and HC). Data to be extracted will differ depending on the type of publication but will include standard bibliographic information, study-specific information for primary studies, and key findings. A preliminary data extraction form is provided in table 1. The draft extraction tool will be piloted on a subset of sources to be included in the review to test its feasibility for the review. The final completed tools will be presented in the scoping review report.

**Table 1.** Data extraction framework.

| **Bibliometrics** | **Characteristics of primary studies** |
| --- | --- |
| Authors | Objectives/ purposes |
| Year of publication | Study location |
| Title | Participants |
| Source | Methodology |
| Country | Key findings |

### Stage 5: Collating, Summarising and Reporting the Results

Levac and colleagues (2010) further break Stage Five into three steps: analyzing the data, reporting results, and applying meaning to the results. Analyzing the data involves presenting a descriptive numerical summary that describe the characteristics of included studies and a thematic analysis that requires qualitative content analytical techniques. The thematic approach described by Braun and Clarke (2006) to capture the broad themes in the data. To report results, the approach that can best stating the outcome will be used. As the scoping review primarily focuses on the environmental features that are found to be influential to residents’ recreational activity experience, which are descriptive in nature, we anticipate using text and graphical approaches to presenting the data, Finally, Levac et al.’s (2010) approach requires researchers to consider the implications of findings for practice, policy and research within the broader context. The authors anticipate the findings will be a critical step in providing evidence-based guidance to inform future practice, policy and research.

## Ethics and Dissemination

Ethical approval and consent to participate are not required for this scoping review. All data generated from the review will be stored on password-protected computers.

The completed scoping review will be submitted for publication in an open-access, peer-reviewed interdisciplinary journal, with the findings disseminated through presentations at regional, national, and international conferences. The results will also be made accessible to health professionals, policymakers, decision-makers, and the public to maximize their impact.

## Data Availability

All data produced in the present work are contained in the manuscript

## Authors’ contributions

ZZ originated the concept, designed the research protocol and methods, and was responsible for drafting and revising the final manuscript. HC helped to refining the research question and study methods and made meaningful contributions to drafting and revising the manuscript. All authors gave their approval for the final version submitted.

## Footnotes

1 Wenjin Wang is not listed as an author in this article as her involvement in this review is only limited to screening of items.

## References

Allen, J. E. (2014). Nursing home federal requirements: Guidelines to surveyors and survey protocols. Springer Publishing Company.

Anderiesen, H., Scherder, E. J., Goossens, R. H., & Sonneveld, M. H. (2014). A systematic review–physical activity in dementia: the influence of the nursing home environment. Applied ergonomics, 45(6), 1678–1686.

Arksey, H., & O’malley, L. (2005). Scoping studies: towards a methodological framework. International journal of social research methodology, 8(1), 19–32.

Barbosa, A., Nolan, M., Sousa, L., & Figueiredo, D. (2014). Dementia in long-term care homes: direct care workers’ difficulties. Procedia-Social and Behavioral Sciences, 140, 172–177.

Beerens, H. C., Zwakhalen, S. M., Verbeek, H., ES Tan, F., Jolani, S., Downs, M., … & Hamers, J. P. (2018). The relation between mood, activity, and interaction in long-term dementia care. Aging & mental health, 22(1), 26–32.

Braun, V., & Clarke, V. (2006). Using thematic analysis in psychology. Qualitative research in psychology, 3(2), 77–101.

British Columbia Ministry of Health. (2024). Long-term care services. Government of British Columbia. https://www2.gov.bc.ca/gov/content/health/accessing-health-care/home-community-care/care-options-and-cost/long-term-care-services

Buettner, L. L., & Fitzsimmons, S. (2003). Activity calendars for older adults with dementia: What you see is not what you get. American Journal of Alzheimer’s Disease & Other Dementias®, 18(4), 215–226.

Calkins, M. P. (2009). Evidence-based long term care design. NeuroRehabilitation, 25(3), 145–154.

Chung, J. C. (2004). Activity participation and well-being of people with dementia in long-term—care settings. OTJR: Occupation, Participation and Health, 24(1), 22–31

Cohen-Mansfield, J., & Werner, P. (1999). Outdoor wandering parks for persons with dementia: a survey of characteristics and use. Alzheimer Disease & Associated Disorders, 13(2), 109–117.

Ducak, K., Denton, M., & Elliot, G. (2018). Implementing Montessori Methods for Dementia™ in Ontario long-term care homes: Recreation staff and multidisciplinary consultants’ perceptions of policy and practice issues. Dementia, 17(1), 5–33.

Edvardsson, D., Petersson, L., Sjogren, K., Lindkvist, M., & Sandman, P. O. (2014). Everyday activities for people with dementia in residential aged care: Associations with personLJcentredness and quality of life. International Journal of Older People Nursing, 9(4), 269–276.

Fleming, R., & Purandare, N. (2010). Long-term care for people with dementia: environmental design guidelines. International psychogeriatrics, 22(7), 1084–1096.

Fortune, D., & Dupuis, S. L. (2018). The potential for leisure to be a key contributor to long-term care culture change. Leisure/Loisir, 42(3), 323–345.

Harmer, B. J., & Orrell, M. (2008). What is meaningful activity for people with dementia living in care homes? A comparison of the views of older people with dementia, staff and family carers. Aging and Mental health, 12(5), 548–558.

Holthe, T., Thorsen, K., & Josephsson, S. (2007). Occupational patterns of people with dementia in residential care: an ethnographic study. Scandinavian journal of occupational therapy, 14(2), 96–107.

Kelly, F. (2010). Recognising and supporting self in dementia: a new way to facilitate a person-centred approach to dementia care. Ageing & Society, 30(1), 103–124.

Klapwijk, M. S., Caljouw, M. A., Pieper, M. J., van der Steen, J. T., & Achterberg, W. P. (2016). Characteristics associated with quality of life in long-term care residents with dementia: a cross-sectional study. Dementia and geriatric cognitive disorders, 42(3-4), 186–197.

Kolanowski, A. M., Buettner, L., Costa, P., & Litaker, M. (2001). Capturing interests: Therapeutic recreation activities for persons with dementia. Therapeutic Recreation Journal, 35(3), 220–235.

Leitner, M. J., & Leitner, S. F. (2004). Leisure in later life (No. Ed. 3). Haworth Press Inc.

Levac, D., Colquhoun, H., & O’brien, K. K. (2010). Scoping studies: advancing the methodology. Implementation science, 5, 1–9.

Lopez, K. J., & Dupuis, S. L. (2014). Exploring meanings and experiences of wellness from residents living in long-term care homes. World Leisure Journal, 56(2), 141–150.

MacDonald, K. C. (2006). Family and staff perceptions of the impact of the long-term care environment on leisure. Topics in Geriatric Rehabilitation, 22(4), 294–308.

Marshall, M. J., & Hutchinson, S. A. (2001). A critique of research on the use of activities with persons with Alzheimer’s disease: A systematic literature review. Journal of advanced nursing, 35(4), 488–496.

Misiorski, S., & Kahn, K. (2006). Changing the culture of long-term care: Moving beyond programmatic change. Journal of Social Work in Long-Term Care, 3(3-4), 137–146.

Narsakka, N., Suhonen, R., Kielo-Viljamaa, E., & Stolt, M. (2022). Physical, social, and symbolic environment related to physical activity of older individuals in long-term care: A mixed-method systematic review. International Journal of Nursing Studies, 135, 104350.

Palacios-Ceña, D., Gómez-Calero, C., Cachón-Pérez, J. M., Velarde-García, J. F., Martínez-Piedrola, R., & Pérez-De-Heredia, M. (2016). Is the experience of meaningful activities understood in nursing homes? A qualitative study. Geriatric Nursing, 37(2), 110–115.

Phinney, A., & Moody, E. M. (2011). Leisure connections: Benefits and challenges of participating in a social recreation group for people with early dementia. Activities, Adaptation & Aging, 35(2), 111–130.

Phinney, A., Chaudhury, H., & O’connor, D. L. (2007). Doing as much as I can do: The meaning of activity for people with dementia. Aging and Mental Health, 11(4), 384–393.

Public Health Agency of Canada. (2019). A dementia strategy for Canada: Together we aspire. https://www.canada.ca/content/dam/phac-aspc/images/services/publications/diseases-conditions/dementia-strategy/National%20Dementia%20Strategy_ENG.pdf

Roland, K. P., & Chappell, N. L. (2015). Meaningful activity for persons with dementia: Family caregiver perspectives. American Journal of Alzheimer’s Disease & Other Dementias®, 30(6), 559–568.

Saperstein, A. R., Calkins, M. P., Van Haitsma, K., & Curyto, K. J. (2004). Missed opportunities: the disconnect between physical design and programming and operations. Alzheimer’s Care Today, 5(4), 324–331.

Schreiner, A. S., Yamamoto, E., & Shiotani, H. (2005). Positive affect among nursing home residents with Alzheimer’s dementia: the effect of recreational activity. Aging & mental health, 9(2), 129–134.

Seetharaman, K., Chaudhury, H., Kary, M., Stewart, J., Lindsay, B., & Hudson, M. (2022). Best practices in dementia care: A review of the grey literature on guidelines for staffing and physical environment in long-term care. Canadian Journal on Aging/La Revue canadienne du vieillissement, 41(1), 55–70.

Smit, D., Willemse, B., de Lange, J., & Pot, A. M. (2014). Wellbeing-enhancing occupation and organizational and environmental contributors in long-term dementia care facilities: an explorative study. International Psychogeriatrics, 26(1), 69–80.

Smit, D., De Lange, J., Willemse, B., Twisk, J., & Pot, A. M. (2016). Activity involvement and quality of life of people at different stages of dementia in long term care facilities. Aging & Mental Health, 20(1), 100–109.

Tak, S. H., Kedia, S., Tongumpun, T. M., & Hong, S. H. (2015). Activity engagement: Perspectives from nursing home residents with dementia. Educational gerontology, 41(3), 182–192.

Tierney, L., MacAndrew, M., Doherty, K., Fielding, E., & Beattie, E. (2023). Characteristics and value of ‘meaningful activity’for people living with dementia in residential aged care facilities:”You’re still part of the world, not just existing”. Dementia, 22(2), 305–327.

Voelkl, J. E., Winkelhake, K., Jeffries, J., & Yoshioka, N. (2003). Examination of Nursing Home Environment: Are Residents Engaged in Recreation Activities?. Therapeutic Recreation Journal, 37(4), 300–314.

Wood, W., Harris, S., Snider, M., & Patchel, S. A. (2005). Activity situations on an Alzheimer’s disease special care unit and resident environmental interaction, time use, and affect. American Journal of Alzheimer’s Disease & Other Dementias®, 20(2), 105–118.

